# Evaluation of COVID-19 as a risk factor for maternal-fetal and neonatal complications: protocol of a systematic review and meta-analysis of cohort and case-control studies

**DOI:** 10.1101/2021.02.23.21252294

**Authors:** Priscila Bezerra, Fernanda Gabriella de Siqueira Barros Nogueira, Alan Chaves dos Santos, Anna katharina Souza Lima, Davi Emanuel Ribeiro, Elias Almeida Silva Barbosa, Suellen Casado dos Santos, Clístenes Crístian de Carvalho

## Abstract

**Background:** COVID-19 in pregnant women has been suggested to impair maternal-fetal and neonatal outcomes. We then designed the present systematic review with meta-analysis to evaluate the repercussion of such disease over maternal fetal and neonatal mortality, need for intensive care, way of delivery, premature delivery, birth weight, Apgar score, presence of intrauterine growth restriction (IGR), and presence of amniotic fluid change.

**Methods:** We will conduct a computerized search through MEDLINE/PubMed, LILACS/BIREME, Web of science, Biorxiv, Medrxiv, and Embase on July 23, 2020. We will include cohort and case-control studies fully reported comparing pregnant women with COVID-19 with those not affected by the disease for maternal fetal and neonatal mortality, need for intensive care, way of delivery, premature delivery occurrence, birth weight, Apgar scores, presence of intrauterine growth restriction, and presence of amniotic fluid change. Three doubles of reviewers will perform in duplicate and independently all steps on screening, risk of bias judgments, and data extraction with ability to discuss disagreements with supervising authors. Pooled effects will be estimated by both fixed and random-effects models and presented according to qualitative and quantitative heterogeneity assessment. Sensitivity analyses will be performed as well as a priori subgroup, meta-regression and multiple meta-regression analyses. We’ll also evaluate the risk of selective publication by assessing funnel plot asymmetry and the quality of the evidence by the application of the GRADE recommendations.

**Discussion:** This systematic review with meta-analysis aims to assess the repercussion of COVID-19 in pregnant women over maternal-fetal and neonatal outcomes and to help clinicians and health systems improve such population outcomes throughout the current pandemic.

**Systematic review registration:** This review protocol was also submitted to PROSPERO registration on February 9, 2021.

## INTRODUCTION

The infection caused by the new coronavirus (SARS-CoV-2) and the subsequent new coronavirus disease 19 (COVID-19) is a global health emergency. Since the first reported cases of an unknown viral pneumonia, in december of 2019, in Wuhan, China, the virus has quickly spread throughout the world, ^1^ being officially declared, by the World Health Organization (WHO), as a pandemic in March of 2020. ^2^ Afterward, there have been 90.335.008 confirmed cases and more than 1.954.336 deaths around the globe.^3^ Thus, this new infection has become the most frequently respiratory infection, which in some cases may evolve to death. ^4^

In january of 2020, chinese scientists had shared the genetic sequence of the new virus, which was later referenced as Severe Acute Respiratory Syndrome Coronavirus 2 (SARS-CoV-2), due to its similarity to the genetic sequence of another virus from the beta-coronavirus family, the SARS-CoV-1, which caused an outbreak worldwide between 2002 and 2003.^3^ Rapidly, there was an increase in the amount of information and knowledge received from chinese researchers about the genetic, virologic, epidemiologic, and clinical aspects of this emergent agent -the seventh coronavirus to be identified as a causer of human infection.^4^

About the clinical presentation, the COVID-19 shows a vast spectrum, once most cases are presented as a mild infectious condition, which symptoms are fever, cough, myalgia, sore throat, headache, digestive disorders, and others. ^2^ However, severe symptoms, as pneumonia and Acute Respiratory Distress Syndrome (ARDS), can also occur, especially in risk groups, e.g., older people (60+ years), pregnant women, immunocompromised patients, and others.^2,5^

In pregnant women, it is paramount to elucidate that they are in a process of characteristic adaptive physiological modifications very common during this phase, such as the alterations in the cardiopulmonary system, i.e., increase in the cardiac frequency, in the systolic volume, and in the oxygen consumption, and, on the contrary, decreasing in lung capacity, as well as the development of an immunosuppressed state.^4,5^

Such particularities can have systemic effects that may increase the risk of developing an acute respiratory disease and its complications, in particular the respiratory insufficiency, which is the reason why this group demands special attention, if infected by the SARS-CoV-2.^6^ In addition to that, there are evidences that healthy pregnant women presents an elevation in the thrombin generation, and intravascular inflammation, promoting a prothrombotic state, which is exacerbated in the context of an infection, thus these patients can also be involvedwith a higher risk of thrombotic events, if affected by the COVID-19. ^7^

In this scenario, it is important to consider data from previous respiratory infectious diseases outbreaks, such as the Severe Acute Respiratory Syndrome (SARS), and the Medium Eastern Respiratory Syndrome (MERS), both caused by coronaviruses (SARS-CoV-1 and MERS-CoV, respectively). Studies reported that these infectious diseases were responsible for an elevated number of maternal complications, such as hospitalization in intensive care, need for assisted ventilation, renal insufficiency, and death. Furthermore, in pregnant women who had SARS and MERS there was a higher number of preterm births, growth restriction, abortion, and fetal death ^6,8^. In a study published by the American Journal of Obstetrics and Gynecology, circa 50% of pregnant women who developed SARS were admitted in the ICU due to a decrease in blood oxygen saturation, about 33% required mechanical ventilation and the mortality rate reached 25% for these women^9.^

Noteworthy, there is a worldwide concern about the risk of intrauterine transmission following a primary maternal infection, as well as how pregnant women react against the SARS-CoV-2 infection. According to clinical studies, when analyzing pregnant women without previous comorbidities and confirmed for SARS-CoV-2 infection, most of the women presented mild to moderate symptoms ^10,11^. However, there is not sufficient evidence for SARS-CoV-2 vertical transmission ^12^.

. Despite the efforts to obtain this knowledge, many key information’s about the SARS-CoV-2 and its effects are still unknown, or limited, including data about pregnant women and their unborn or newborn baby, which difficult the basis for specific care recommendations during the pregnancy. ^13,14^ Therefore, it is not totally clear whether the clinical characteristics of pregnant women affected by the virus differs from non-pregnant women or not, if parturition and puerperium worsen the respiratory symptoms of COVID-19, then leading to an increase of the maternal and fetal morbimortality, in addition to questioning about the possible fetal alterations, i.e., early premature labor, decreasing of amniotic liquid, acute fetal distress, intrauterine growth restriction, and others.^15^

As stated above, it is satisfactory to seek, identify and search in medical literature information about the relation of COVID-19 and pregnant women, aiming to contribute with the scientific community in the clinical management and treatment of this key group of patients.

## METHODS

### Protocol and registration

The current protocol was designed according to recommended standards and reported as per the Preferred Reporting Items for Systematic Reviews and Meta-analysis Protocols (PRISMA-P) guidelines^16^. This review protocol was also submitted to PROSPERO registration on February 9, 2021.

### Eligibility criteria

Inclusion criteria will be as follows: 1) cohort and case-control studies fully reported; 2) pregnant patient 3) data available on maternal fetal and neonatal mortality, need for intensive care, way of delivery, premature delivery, birth weight, Apgar score, presence of intrauterine growth restriction (IGR), and presence of amniotic fluid change. 4) Comparison between pregnant women with and without COVID-19. Exclusion criteria will be as follows: 1) studies published in language other than English, Spanish or Portuguese; 2) inability to abstract outcome data at patient level.

### Information sources

We will conduct a computerized search through MEDLINE/PubMed, LILACS/BIREME, Web of science, Biorxiv, Medrxiv, and Embase on July 23, 2020. We will also search the reference lists of included studies.

### Search strategy

The following searching strategy line will be applied to databases with no limitations: COVID-19[Supplementary Concept] OR “coronavirus disease” OR covid19 OR covid-19* OR SARS-CoV-2 OR “SARS-CoV-2 infection” OR “2019 novel coronavirus” OR 2019-nCoV OR “COVID-19 virus disease” OR “coronavirus disease 2019” OR “coronavirus disease-19” OR “2019-nCoV disease” OR “COVID-19 virus infection” OR “severe acute respiratory syndrome coronavirus 2”[Supplementary Concept] OR “wuhancoronavirus” OR “COVID-19 virus” OR “coronavirus disease 2019 virus” OR “COVID19 virus” OR “wuhanseafood market pneumonia virus”) AND (fetus[MeSH] OR fetus* OR “fetal structure*” OR fetal OR “fetal tissue” OR “retained fetus” OR pregnancy[MeSH] OR pregnanc* OR gestation OR “pregnant women”[MeSH] OR “prenatal care”[MeSH] OR “antenatal care” OR “infant, newborn”[MeSH] OR newborn* OR neonate* OR “newborn infant*” OR “pregnancy, high-risk”[MeSH] OR “high-risk pregnanc*” OR “high risk pregnanc*” OR “pregnancy complications”[MeSH] OR “complication, pregnancy” OR “complications, pregnancy” OR “pregnancy outcome”[MeSH] OR “pregnancy outcomes” OR “outcome, pregnancy” OR “outcomes, pregnancy” OR “maternal mortality”[MeSH] OR “maternal mortalit*” OR “mortality, maternal” OR “infant, newborn, diseases”[MeSH] OR “neonatal disease*” OR “disease, neonatal” OR “diseases, neonatal” OR “congenital abnormalities”[MeSH] OR “congenital abnormalit*” OR “congenital defects” OR “birth defect*” OR deformit* OR “fetal distress”[MeSH] OR “nonreassuring fetal status” OR “fetal diseases”[MeSH] OR “fetal disease*” OR “embryopath*” OR “perinatal death”[MeSH] OR “perinatal death*” OR “neonatal death*” OR “death, perinatal” OR “deaths, perinatal” OR “death, neonatal” OR “deaths, neonatal”)

### Study records

Retrieved references will be taken to EPPI Reviewer Web (Beta) for screening steps – “title and abstract” then “full text”. Eligibility criteria will be applied to select the studies to be included. Three doubles of reviewers will perform in duplicate and independently all steps from screening of title and abstract, through screening of full text and risk of bias assessment to data extraction. The results will be compared, and disagreements solved by discussion and consensus amongst correspondent researchers and PB. In case of no consensus, AC will act as final judge. In case of missing data, corresponding authors will be reached. If relevant data is missing or data are conflicting and corresponding author do not reply our contact after three attempts, then the article will be excluded. Data will be recorded in an excel spreadsheet. The data-extraction form will first be tested in 5 included studies and then refined if necessary.

### Data items

We will collect data on first authors name, publication year, study design, characteristics of population and country. Also, we will collect maternal date such as mean age, mean BMI, mean weight, mean height, mean gestational age, presence of preview maternal comorbidity (diabetes, respiratory disease, hypertension and thrombophilia), number of patients enrolled and analyzed of each study, number of participants in each arm. Also, for continuous outcomes, number of patients, mean and standard deviation in each arm and mean difference and standard error between groups. For categorical outcomes, number of events and number of patients in each arm along with odds ratio and standard error between groups.

### Outcomes with prioritization

We will primarily investigate maternal-fetal and neonatal mortality – which authors consider the most important outcomes when assessing risk factors for maternal-fetal and neonatal complications. To getting further information on COVID-19 rebounds over maternal-fetal and neonatal health, we will also evaluate need for intensive care, way of delivery (normal or cesarean delivery), preterm delivery occurrence (before 37 weeks of pregnancy are completed) mean birth weight or birth weight status (low birth weight - less than 2,500 g; Very low birth weight - less than 1500 g; and extremely low birth weight - less than 1 000 g), Apgar scores at 1^st^ and 5^th^ minute, presence of IGR (fetus weighing less than the 10th percentile for gestational age), and presence of amniotic fluid change (normal values being those above the 2.5 percentile of the amniotic fluid index curve).

### Risk of bias in individual studies

We’ll apply the “Quality in Prognosis Studies” (QUIPS)^17^ tool that has been developed to assess the study quality and risk of bias (RoB) and consists of several prompting items categorized into six domains (Study participation, Study attrition, Prognostic factor measurement, Outcome measurement, Study confounding, Statistical analysis and reporting). Overall, each domain is going to be judged on a three-grade scale (low, moderate or high risk of bias). This categorization is based on the following criteria: If all domains are classified as having low RoB, or up to one moderate RoB, then this paper will be classified as low RoB. If one or more domains are classified as having high RoB, or ≥ 3 moderate RoB, then this paper will be classified as high RoB. All papers in between will beclassified as having moderate RoB. This categorization is going to be a result of continuous discussion between the authors. We will also use risk of bias judgments for sensitivity analysis.

### Data synthesis

Data will be summarized if at least two different sources are available. Analyses will be conducted in Review Manager (RevMan, London, UK, v5.3.5) and R software tools (R Foundation for Statistical Computing, Vienna, Austria), as appropriate. Data on patient level will be extracted or calculated from studies and used for summarizations. Effect sizes, SE, and 95% CI will be estimated for each study from the recorded data. Forest plots will be constructed for every outcome. Pooled estimates will be calculated by both fixed-effects (Mantel-Haenszel or inverse variance method, where appropriate) and random-effects (Sidik-Jonkman method with Hartung-Knapp adjustment) for sensitivity analyses. Heterogeneity will be evaluated qualitatively and quantitatively by Cochran’s Q-test and *I*^*2*^. Where qualitative or quantitative heterogeneity is present, pooled estimates from random-effects models will be presented. An influence analysis by Leave-One-Out method will be performed to assess the influence of each study in the pooled effects and the between studies heterogeneity. Sensitivity analyses will be undertaken throughout both subgroup and meta-regression analyses. Subgroup assessments will be performed using either mixed-effects or random-effects models, where appropriate, for outcomes with 10 or more studies available. Meta-regressions of single features, one at a time, as well as multiple meta-regression with maximum likelihood (ML) estimator for *τ2* will be conducted only for outcomes with 10 or more studies available per covariate. The multiple meta-regression models will be submitted to a permutation test to confirm statistical significance. Both subgroup and meta-regression analyses will be performed with a priori hypotheses – attempting to avert the catch of spurious associations – with the following features: intrauterine growth restriction, change in amniotic fluid, premature rupture of membranes, intrauterine fetal death, premature labor, and maternal clinical changes, such as diabetes, hypertensive syndromes, coagulopathies, lung diseases and thromboembolic events. Risk of bias judgments and decisions made throughout the statistical analysis will also be included in sensitivity analyses.

### Meta-bias

We will perform assessment of selective publication by small sample bias methods for those outcomes with 10 or more studies. Funnel plots will be built, and Egger’s tests performed to check for plot asymmetry. The threshold of significance will be set at p<0.100 for this method as this test has low power. Where asymmetries are present, a Duval and Tweedie’s trim-and-fill procedure will be applied to estimate bias-corrected effects.

### Confidence in cumulative evidence

To assess the quality of the evidence, we will apply the Grading of Recommendations, Assessment, Development and Evaluation (GRADE) approach. This approach takes into account factors related to design of the study, risk of bias, inconsistent results, indirectness of evidence, imprecision, publication bias, the magnitude of the effect, existence of dose-response gradient, and if all plausible confoundings would only reduce the demonstrated effect.

## DISCUSSION

Some studies have suggested ACE2 from pregnant women to be deregulated by SARS-CoV-2, potentially increasing the risk of pre-eclampsia, as well as fetal nutrition to be impaired, also increasing chances of intrauterine growth restriction. We then designed the present systematic review with meta-analysis to analyze the repercussion of COVID-19 in pregnant women over maternal-fetal and neonatal outcomes.^6,12,13^ We aim to help clinicians and health systems to make evidence-based decisions in order to improve this population outcomes throughout the pandemic.

## Data Availability

We conduct the protocol according to prisma P and submitted the protocol to registration in prospero.

## CONTRIBUTIONS

### Design of the project

Priscila Bezerra, Fernanda Gabriella de Siqueira Barros Nogueira, Alan chaves dos santos, Anna katharina Souza Lima, Davi Emanuel ribeiro, Elias Barbosa Almeida da Silva, Suellen casado dos santos, Clístenes Crístian de Carvalho.

### Writing of the protocol

Priscila Bezerra, Fernanda Gabriella de Siqueira Barros Nogueira, Alan chaves dos santos, Clístenes Crístian de Carvalho

### Review of the protocol

Priscila Bezerra, Fernanda Gabriella de Siqueira Barros Nogueira, Alan chaves dos santos, Clístenes Crístian de Carvalho.

### Final protocol approval

Priscila Bezerra, Fernanda Gabriella de Siqueira Barros Nogueira, Alan chaves dos santos, Anna katharina Souza Lima, Davi Emanuel ribeiro, Elias Barbosa Almeida da Silva, Suellen casado dos santos, Clístenes Crístian de Carvalho.

